# A Multiplex Serological Assay for Quantification of Arbovirus Antibody Kinetics and Neutralizing Antibodies

**DOI:** 10.1101/2025.10.31.25339243

**Authors:** Laura Garcia, Victor Yman, Marie-Fabrice Gasasira, Françoise Donnadieu, Lucie Cappuccio, Damien Hoinard, Philippe Desprès, Jean-Claude Manuguerra, In-Kyu Yoon, Taweewan Hunsawong, Maria Theresa P Alera, Emily S. Gurley, Nikos Vasilakis, Scott C. Weaver, Ziaur Rahman, Ilaria Dorigatti, Chris Drakeley, Inès Vigan-Womas, Makhtar Niang, Henrik Salje, Jessica Vanhomwegen, Michael White

## Abstract

Arboviruses such as dengue virus (DENV) and chikungunya virus (CHIKV) pose major global health threats. To support serological surveillance and study cross-reactivity, an in-house multiplex bead-based immunoassay was developed to measure IgG responses against a wide panel of orthoflavivirus and alphavirus antigens. Samples from Senegal, France, the Philippines, and Bangladesh enabled analyses to be carried out in endemic and non-endemic regions. IgG kinetics following DENV1 and CHIKV infections revealed virus-specific temporal profiles and identified antigens, such as CHIKV E2 and VLP, as well as DENV1 EDIII and SNAP-tagged EDIII, as promising markers for serosurveillance. A two-step Random Forest model was used to classify PRNT-positive samples and predict PRNT titers. CHIKV showed a strong correlation between IgG and PRNT titers (R^2^ = 0.71), while DENV1 showed weaker performance (R^2^ = 0.42). These results support the use of multiplex serology for arbovirus surveillance and highlight the limitations of using binding antibodies to predict neutralisation, particularly for orthoflaviviruses.

## Introduction

Arthropod-borne viruses (arboviruses) are transmitted to humans and other vertebrates by blood-feeding arthropods, such as mosquitoes, flies, or ticks. Arboviruses of most human impact are primarly spread by *Aedes spp.* mosquitoes, particularly *Aedes aegypti* and *Aedes albopictus*(1). Globally, more than 500 arboviruses have been identified, of which 150 are known to cause disease in humans. Arboviruses belong to several families, including *Flaviviridae* (e.g., the orthoflaviviruses dengue (DENV), Zika (ZIKV), yellow fever (YFV), Usutu (USUV), West Nile (WNV), japanese encephalitis viruses (JEV)), and *Togaviridae* (e.g., the *alphaviruses* chikungunya (CHIKV), Mayaro (MAYV), Ross River (RRV), and O’nyong nyong (ONNV) viruses). Dengue virus remains the most prevalent, with 14 millions cases and 10,000 deaths reported in 2024(2). Chikungunya virus caused 480,000 reported cases and 200 reported deaths in the same year(3).

Although the major Zika virus epidemic occurred in 2016(4), particularly in Brazil(5) and over 50 other countries worldwide, sporadic and localized cases continue to be reported, indicating a persistent but reduced public health threat(6).

Understanding the circulation of arboviruses within human populations is essential for guiding public health interventions, informing vaccine development and vaccination strategies, and enhancing outbreak preparedness. Seroprevalence studies, which assess the presence of virus-specific antibodies, provide crucial insights into past exposure, immunity levels and transmission dynamics. These studies play a key role in estimating the true burden of arboviral infections, identifying high-risk populations, and assessing the potential for future outbreaks. However, conducting seroprevalence studies presents several challenges, particularly in regions where multiple arboviruses co-circulate(7).

One of the main challenges in arbovirus serology is the high degree of antibody cross-reactivity among orthoflaviviruses. Due to their structural similarities, antibodies elicited by one virus, such as dengue virus, can cross-react with another, such as Zika or West Nile virus, leading to false-positive results and complicating accurate serodiagnosis. Furthermore, original antigenic sin may further influence immune responses, as prior exposure to one orthoflavivirus affects subsequent responses to related viruses, including memory-driven increases in antibody levels to the first related virus to infect a person(8,9). Although some degree of cross-reactivity can occur among other arbovirus groups, such as alphaviruses, it is typically more limited(10).

Traditional serological assays, including enzyme-linked immunosorbent assays (ELISA), remain widely used for detecting arboviral antibodies. IgM capture ELISA is typically employed for early infection detection, while indirect IgG assays are used to assess long-term immune responses(11). However, these assays often lack specificity, particularly in endemic regions with multiple co-circulating arboviruses. Other approaches, such as plaque reduction neutralization tests (PRNTs), often offer greater specificity by measuring virus-neutralizing antibodies. Despite their reliability, PRNTs are labor-intensive, and sometimes require biosafety level 3 (BSL-3) containment, limiting their accessibility, particularly in resource-limited settings(12). Rapid diagnostic tests (RDTs) have emerged as potential alternatives, but their sensitivity and specificity remain variable, necessitating further validation for widespread implementation(13).

There is a critical need for improved serosurveillance tools that offer greater specificity, sensitivity, and scalability. Multiplex bead-based assays have emerged as a promising option for arbovirus serology, addressing many of the limitations of conventional techniques(14),(15,16). Unlike conventionnal ELISA or PRNT, multiplex assays enable the simultaneous detection of antibodies against multiple pathogens in a single reaction, significantly reducing the time, labor, and costs associated with testing as well as the volumes of serum or plasma needed. This high-throughput approach enhances the ability to reconstruct exposure history, making it particularly valuable in regions where multiple arboviruses co-circulate. Additionally, by using carrefully selected/ engineered recombinant viral antigens and optimized coupling strategies, multiplex assays can improve specificity and minimize cross-reactivity, thereby increasing the reliability of seroprevalence studies. The development and implementation of such advanced serological tools are essential for strengthening arbovirus surveillance and improving outbreak preparedness worldwide.

## Materials and Methods

### Samples

This study utilized panels of samples collected from multiple countries (Table 1).

**Table 1:** Overview of participant and sample data, stratified by country.

| country | expected arbovirus exposure | number of participants | number of samples | median age [range] | male (%) | female (%) |
| --- | --- | --- | --- | --- | --- | --- |
| Bangladesh | DENV, JEV and CHIKV | 78 | 78 | 26.5 [6–84] | 52.6 | 47.4 |
| France | non-endemic (control) | 84 | 84 | 82.5 [8–101] | 19 | 81 |
| Philippines | DENV, ZIKV and CHIKV | 36 | 215 | 9.5 (1-49) | 49 | 51 |
| Senegal | DENV, ZIKV, YFV, WNV, CHIKV and ONNV | 270 | 270 | 11.2 [0.3–77] | 43.7 | 51.9 |

#### Cross sectional studies

We analyzed a subset of 74 samples from a cross-sectional seroprevalence study conducted in 2014 in the Chapai Nawabganj district (called zila) of Bangladesh, located in the Rajshahi division. These samples were a subset of 1455 samples collected from 40 communities in the district. In this subset, DENV and CHIKV PRNT testing was conducted in addition to multiplex testing(17).

The Senegalese villages of Dielmo and Ndiop have been monitored longitudinally since the 1990’s with repeat cross-sectional studies to investigate malaria epidemiology(18). We analysed 270 samples from cross-sectional seroprevalence studies in these villages with samples collected in 2016 and 2018.

#### Longitudinal study

We analyzed 215 samples from 36 individuals from a longitudinal cohort study conducted in the village of Punta Princesa, located in the Visayas region of the Philippines(15,16). Briefly, study participants had annual study visits including blood collection at baseline, 12 months, and 24 months. Active fever surveillance was conducted through weekly face-to-face or telephone contacts; any history of fever identified triggered an acute and 3-week convalescent visit with blood collection. Acute serum samples were tested by two-step nested reverse transcriptase PCR (RT-PCR) to detect CHIKV, DENV 1 to 4 or ZIKV RNA. All samples from individuals that had a RT-PCR confirmed symptomatic ZIKV (N=1), CHIKV (N=12), DENV1 (N=12), DENV2 (N=6), DENV3 (N=2) and DENV4 (N=3) infection were selected for multiplex testing. This allowed us to track longitudinal serological patterns in PCR confirmed infections.

#### Negative controls

As negative controls, we also analyzed 84 samples from France, obtained from a longitudinal study on SARS-CoV-2 conducted in Crépy-en-Valois in 2021(19).

### Ethics statement

The Bangladesh study was approved by the ethical review board at icddr,b (protocol #PR-14058) and Institut Pasteur (protocol #2015-074). The Senegalese samples were collected as part of the ongoing Dielmo and Ndiop project which was approved by the Senegalese National Health Research Ethics Committee (CNERS Sénégal). Approval to measure antibodies in these samples was granted by CNERS Sénégal (N 00000007 MSAS/CNERS/Sec). The Philippines study was approved by the ethical review boards at Vicente Sotto Memorial Medical Center in Cebu City, Philippines and Walter Reed Army Institute of Research Human Subjects Protection Branch (protocol #1833). Negative samples were obtained from the COVID-Oise study, which was registered with ClinicalTrials.gov (NCT04644159) and received ethical approval from the Comité de Protection des Personnes Nord Ouest IV. Formal consent was obtained from all study participants, or their parents/guardians.

### Antigens

We developed a multiplex bead-based assay comprising 44 antigens (Table 2) from commonly circulating arboviruses, including dengue, Zika, and chikungunya. Recombinant proteins were sourced from Native Antigen Company (Oxford, UK), Cusabio (Houston, USA), Alpha Diagnostics Intl. Inc. (San Antonio, USA) or Clinisciences (Nanterre, France).

**Table 2:** List of antigen names and suppliers used for multiplex bead-based assay development.

| Antigen name | Short name | Strain/ Isolate Information | Supplier | Family | Catalogue number |
| --- | --- | --- | --- | --- | --- |
| Chikungunya soluble ectodomain E2 | CHIKV E2 | Senegal 37997 | Native antigen | *Togaviridae* | REC31617-100 |
| Chikungunya virus-like particles | CHIKV VLP | Senegal 37997 | Native antigen | *Togaviridae* | CHIKV-VLP-100 |
| Chikungunya non structural protein | CHIKV NSP | strain S27-African prototype | Cusabio | *Togaviridae* | CSB-EP810351CJAT |
| SNAP-chikungunya soluble ectodomain E2 | CHIKV E2 + SNAP | 06-049 | In-house production – EVAg | *Togaviridae* | 014P-03540 |
| Dengue 1 domain III envelope protein | DENV1 EDIII | Dengue virus 1 Nauru/West Pac/1974 | Native antigen | *Flaviviridae* | REC32042-100 |
| Dengue 1 non structural protein 1 | DENV1 NS1 | Nauru/Western Pacific/1974 | Native antigen | *Flaviviridae* | DENV1-NS1-100 |
| Dengue 1 virus-like particles | DENV1 VLP | Puerto Rico/US/BID-V853/1998 | Native antigen | *Flaviviridae* | DENVX4-VLP-100 |
| Dengue 2 domain III envelope protein | DENV2 EDIII | Dengue virus 2 Jamaica/1409/1983 | Native antigen | *Flaviviridae* | REC32043-100 |
| Dengue 2 non structural protein 1 | DENV2 NS1 | Thailand/16681/84 | Native antigen | *Flaviviridae* | DENV2-NS1-100 |
| Dengue 2 virus-like particles | DENV2 VLP | Thailand/16681/84. | Native antigen | *Flaviviridae* | DENVX4-VLP-100 |
| Dengue 3 domain III envelope protein | DENV3 EDIII | Dengue virus 3 isolate 15/1329 | Native antigen | *Flaviviridae* | REC32044-100 |
| Dengue 3 non structural protein 1 | DENV3 NS1 | Sri Lanka D3/H/IMTSSA-SRI/2000/1266 | Native antigen | *Flaviviridae* | DENV3-NS1-100 |
| Dengue 3 virus-like particles | DENV3 VLP | Sri Lanka D3/H/IMTSSA-SRI/2000/1266 | Native antigen | *Flaviviridae* | DENVX4-VLP-100 |
| Dengue 4 domain III envelope protein | DENV4 EDIII | DENV-4/SG/06K2270DK1/2005 | Native antigen | *Flaviviridae* | REC32045-100 |
| Dengue 4 non structural protein 1 | DENV4 NS1 | Dominica/814669/1981 | Native antigen | *Flaviviridae* | DENV4-NS1-100 |
| Dengue 4 virus-like particles | DENV4 VLP | Dominica/814669/1981 | Native antigen | *Flaviviridae* | DENVX4-VLP-100 |
| SNAP-Dengue 1 domain III envelope protein | DENV1 EDIII + SNAP | FGA/NA d1d | In-house production – EVAg | *Flaviviridae* | 014P-03514 |
| SNAP-Dengue 2 domain III envelope protein | DENV2 EDIII + SNAP | CS85-3 | In-house production – EVAg | *Flaviviridae* | 014P-03515 |
| SNAP-Dengue 3 domain III envelope protein | DENV3 EDIII + SNAP | DENV-3/TH/BID-V2329/2001 | In-house production – EVAg | *Flaviviridae* | 014P-03516 |
| SNAP-Dengue 4 domain III envelope protein | DENV4 EDIII + SNAP | CS101-1 | In-house production – EVAg | *Flaviviridae* | 014P-03517 |
| Japanese encephalitis envelope protein | JEV E | SA-14 | Native antigen | *Flaviviridae* | REC31688-100 |
| Japanese encephalitis non structural protein 1 | JEV NS1 | SA-14 | Native antigen | *Flaviviridae* | JEV-NS1-100 |
| SNAP Japanese encephalitis domain III envelope protein | JEV EDIII + SNAP | GP05 | In-house production – EVAg | *Flaviviridae* | 014P-03521 |
| Mayaro non structural protein 1 | MAYV NSP | Strain Brazil | Alpha Diagnostics Intl. Inc. | *Togaviridae* | AD-MAYV41-R-25 |
| Mayaro soluble ectodomain E2 | MAYV E2 | Acre27 | Native antigen | *Togaviridae* | REC31644-100 |
| SNAP-Mayaro soluble ectodomain E2 | MAYV E2 + SNAP | Brazil BeH256 | In-house production – EVAg | *Togaviridae* | 014P-03542 |
| O'nyong nyong soluble ectodomain E2 | ONNV E2 | SG650 | Clinisciences | *Togaviridae* | IT-022-006Ep |
| O'nyong nyong virus-like particles | ONNV VLP | SG650 | Native antigen | *Togaviridae* | REC31626-100 |
| Ross River structural polyprotein | RRV SP | T48 | Native antigen | *Togaviridae* | REC31969-100 |
| SNAP Ross River soluble ectodomain E2 | RRV E2 + SNAP | QML1 | In-house production – EVAg | *Togaviridae* | 014P-03541 |
| Rift Valley fever nucleoprotein | RVFV NP | ZH-548 | Native antigen | *Phenuiviridae* | REC31640-100 |
| Usutu non structural protein 1 | USUV NS1 | Vienna 2001 | Native antigen | *Flaviviridae* | USUV-NS1-100 |
| SNAP Usutu domain III envelope protein | USUV EDIII + SNAP | Usu973 | In-house production – EVAg | *Flaviviridae* | 017P-06227 |
| West Nile domain III envelope protein | WNV EDIII | lineage 1, strain NY99 | Sino Biological | *Flaviviridae* | 40345-V08Y |
| West Nile non structural protein 1 | WNV NS1 | NY99 | Native antigen | *Flaviviridae* | WNV-NS1-100 |
| SNAP West Nile domain III envelope protein | WNV EDIII + SNAP | DES 1476-01 | In-house production – EVAg | *Flaviviridae* | 014P-03519 |
| Yellow fever envelope protein | YFV E | 17D-204-USA vaccine, isolate HONG3 | Native antigen | *Flaviviridae* | REC32081-100 |
| Yellow fever non structural protein 1 | YFV NS1 | 17D | Native antigen | *Flaviviridae* | YFV-NS1-100 |
| SNAP Yellow fever domain III envelope protein | YFV EDIII + SNAP | Asibi | In-house production – EVAg | *Flaviviridae* | 014P-03520 |
| Zika Uganda strain non structural protein 1 | ZIKV NS1 | Uganda MR 769 | Native antigen | *Flaviviridae* | ZIKV-NS1-100 |
| Zika Asian strain domain III envelope protein | ZIKVAS EDIII | Zika virus isolate Z582-16 (Brazil, March 2018) | Native antigen | *Flaviviridae* | REC32046-100 |
| Zika Suriname strain non structural protein 1 | ZIKVSU NS1 | Suriname Z1106033 | Native antigen | *Flaviviridae* | ZIKVSU-NS1-100 |
| SNAP-Zika domain III envelope protein | ZIKV EDIII + SNAP | EC YAP | In-house production – EVAg | *Flaviviridae* | 014P-03522 |
| SNAP control | SNAP | - | In-house production – EVAg | *Control* | - |

In parallel, we utilized recombinant orthoflavivirus envelope domain III (EDIII) and soluble ectodomains of alphavirus envelope (E2) protein prepared at Institut Pasteur(20) (Table 2). These recombinant viral antigens were expressed as chimeric proteins fused to a modified O6-alkylguanine-DNA alkyltransferase enzyme (AGT), referred to as the SNAP domain. The SNAP-antigen fusion proteins were coupled to the microspheres using an oriented, two-step coupling procedure to ensure optimal antigen presentation.

### Manual oriented coupling of in-house recombinant proteins

Manual coupling of recombinant proteins containing the SNAP-tag was performed using the Bio-Plex Amine Coupling Kit (Bio-Rad). The first step consists of coupling the AGT substrate BG-PEG-NH_2_ to the activated microspheres by amine coupling, and the second step consists of binding the SNAP-tagged proteins to the substrate-coated microspheres.

The stock of uncoupled microspheres were sonicated and vortexed for 30 seconds. Subsequently, 2.5 × 10⁶ microspheres (200 µL) were transferred to an Eppendorf tube and washed once with bead wash buffer using a magnetic separator. The beads were then activated for 20 minutes at room temperature (RT) with 80 µL of bead activation buffer, 10 µL of hydroxysulfosuccinimide sodium salt (S-NHS; 50 mg/mL, Sigma-Aldrich), and 10 µL of 1-ethyl-3-(3-dimethylaminopropyl)carbodiimide hydrochloride (EDAC; 50 mg/mL, Sigma-Aldrich). Both S-NHS and EDAC were prepared in bead activation buffer immediately prior to use.

Following activation, the beads were washed twice with Phosphate Buffer Saline buffer (PBS), and 5 mg/mL of BG-PEG-NH_2_ was added to the coupling reaction. The beads were incubated overnight at RT on a shaker. After incubation, the beads were washed twice with PBS buffer and resuspended in 250 µL of blocking buffer. This suspension was incubated for 30 minutes in the dark on a shaker.

After blocking, the beads were washed twice with PBS, and 50–100 µg of recombinant proteins was added in 1 mL of PBS. The optimal amount of recombinant proteins (µg) per 1.25x10^6^ of beads was optimised in a previous study, using an anti-SNAP polyclonal antibody (New England Biolabs) to obtain a saturation of the substrate BG-PEG-NH_2_ immobilised on the surface of the microspheres. The suspension was incubated in the dark on a roller for 2 hours. The coupled beads were then washed twice with PBS and resuspended in 1 mL of storage buffer. The beads were stored at 4°C.

One day after the coupling process, the antigen-coupled beads were verified by using an anti-SNAP polyclonal antibody, and the results were compared with those from the optimisation process to confirm coupling efficiency. After verification, all coupled beads were counted using the TC20 Automated Cell Counter (Bio-Rad).

### Automated coupling procedure

For recombinant proteins that do not contain the SNAP-tag, the coupling procedure was automated using the KingFisher Duo Prime magnetic particle processor (Thermo Fisher Scientific, Waltham, MA, USA). This instrument is equipped with a 12-well magnet head and 96-deep well plates. The protocol implemented in the processor was adapted from the manual coupling procedure(21). A detailed description of the BindIt software protocol (version 4.0.0.45) is provided in the supplementary data (Supplementary Methods, Table S1).

This automated method allowed for the coupling of up to 12 recombinant proteins in a single run. Buffers, volumes, and incubation times were adjusted to fit the automated workflow. For each run, the instrument was set up with a bead comb and two deep-well plates (Thermo Fisher Scientific) containing the necessary buffers and microspheres, as specified by the program. The composition and layout of the two plates are detailed in the supplementary methods.

Once the automated run was completed, the coupled beads were transferred from the 96-well plate to Eppendorf tubes. The beads were stored at 4°C, and one day after the coupling process, the antigen-coupled beads were verified using a pool of serum from arbovirus-exposed individuals to assess whether they generated a log-linear standard curve. Then, the coupled beads were counted using the TC20 Automated Cell Counter (Bio-Rad).

### Multiplex immunoassay (MIA)

The bead-based multiplex immunoassay (MIA) procedure was performed as previously described(22) with some adjustments. Briefly, a mixture of 44 antigen-coupled bead regions (1,000 beads/region/well) was sequentially incubated in the dark under constant shaking with diluted serum samples: 1:400 for IgG and 1:200 for IgA and IgM. The incubation was carried out for 30 minutes on a shaker. All serum dilutions and bead premixes were prepared in PBT buffer (phosphate-buffered saline containing 1% bovine serum albumin and 0.05% (v/v) Tween-20).

Following incubation, the plates were washed three times with PBS 1X containing 0.05% Tween-20 using a HydroSpeed™ plate washer (Tecan). After washing, the plates were incubated for 15 minutes with a secondary antibody conjugated to R-phycoerythrin (2 µg/mL) specific for IgG, IgA, or IgM (Jackson Immunoresearch, UK). The plates were then washed three more times using the HydroSpeed™ plate washer and resuspended in 150 µL of PBS 1X with 0.05% Tween-20. Plates were read using the Intelliflex® system at a low Photomultiplier Tube (PMT) setting, and the median fluorescence intensity (MFI) was measured.

Samples were run on 96 well plates, containing a blank (only beads, without serum) and a standard curve prepared from two-fold serial dilutions (1:50 to 1:102 400) of a pool of serum from arbovirus-exposed individuals. A normalised MFI (nMFI) for the antibody response to an antigen in a sample was calculated by dividing the measured MFI by the MFI of the 1:400 dilution of the pool of serum from arbovirus-exposed individuals. For antigens containing a SNAP tag, the nMFI was calculated by dividing the measured MFI in a sample by the MFI to the SNAP tag only in the same sample.

The protocol for measuring IgG avidity is detailed in the Supplementary Methods.

Analyses were performed using R (version 4.4.3).

### Plaque reduction neutralisation test (PRNT)

Plaque reduction neutralisation tests (PRNT) were performed using the procedure of Russell et *al.*(23) and others(24). In brief, four-fold serial dilutions of sera from 1:10 were incubated with attenuated CHIKV strain 181/clone25 derived from a CHIKV Asian strain isolated from Thailand in 1962(25). Mixtures of virus-serum were incubated on a monolayer of Macaca mulatta kidney (LLC-MK2) cells in a 12-well plate with 30–50 plaques in control wells. PRNT titers were expressed as the extrapolated serum dilution causing 80% plaque reduction. DENV PRNTs in Bangladesh were conducted by the Vasilakis laboratory at UTMB as described previously(26).

### Random forest models for predicting arboviral PRNTs using multiplex serological data

We used a two-step random forest modelling approach to assess whether the multiplex serological data could be used to predict PRNT titers for DENV (Serotype 1-3, Supplementary Figures S11 and S16) and CHIKV in Bangladesh. We did not fit models to DENV serotype 4 because of low rates of positivity in the tested samples. The model was fitted separately to PRNT data for each DENV serotype and CHIKV and cross-validation was used to tune random forest model hyperparameters at each step. Model performance was assessed using out-of-bag (OOB) predictions, which serve as an internal form of cross-validation.

In step 1 (Classification), a Boruta feature selection algorithm(27) was used to identify all serological responses that contribute significant information for accurate classification of PRNT status (i.e. positive or negative). A random forest classifier was then fitted to the reduced set of serological predictors to determine the PRNT status of each sample.

Step 2 (Regression) was applied to samples that were predicted to be PRNT positive in step 1. In this step, a second Boruta feature selection was performed to identify all serological responses that contribute significant information to predicting the PRNT titers. A random forest regression model was then fitted to the reduced set of predictors to estimate the PRNT titer in PRNT positive samples.

Predictions from both steps were aggregated to obtain a final PRNT titer estimate for each sample which was compared with the actual observed PRNT titer to assess model performance.

## Results

### Distribution of IgG antibody for alphaviruses accross four countries

Figure 1 illustrates the distribution of IgG nMFI against alphavirus antigens from our 44-plex assay in a total of 647 samples from four countries (France, Bangladesh, Senegal and the Philippines). Overall, serological responses varied across countries, suggesting differences in exposure to alphaviruses.

**Figure 1:**
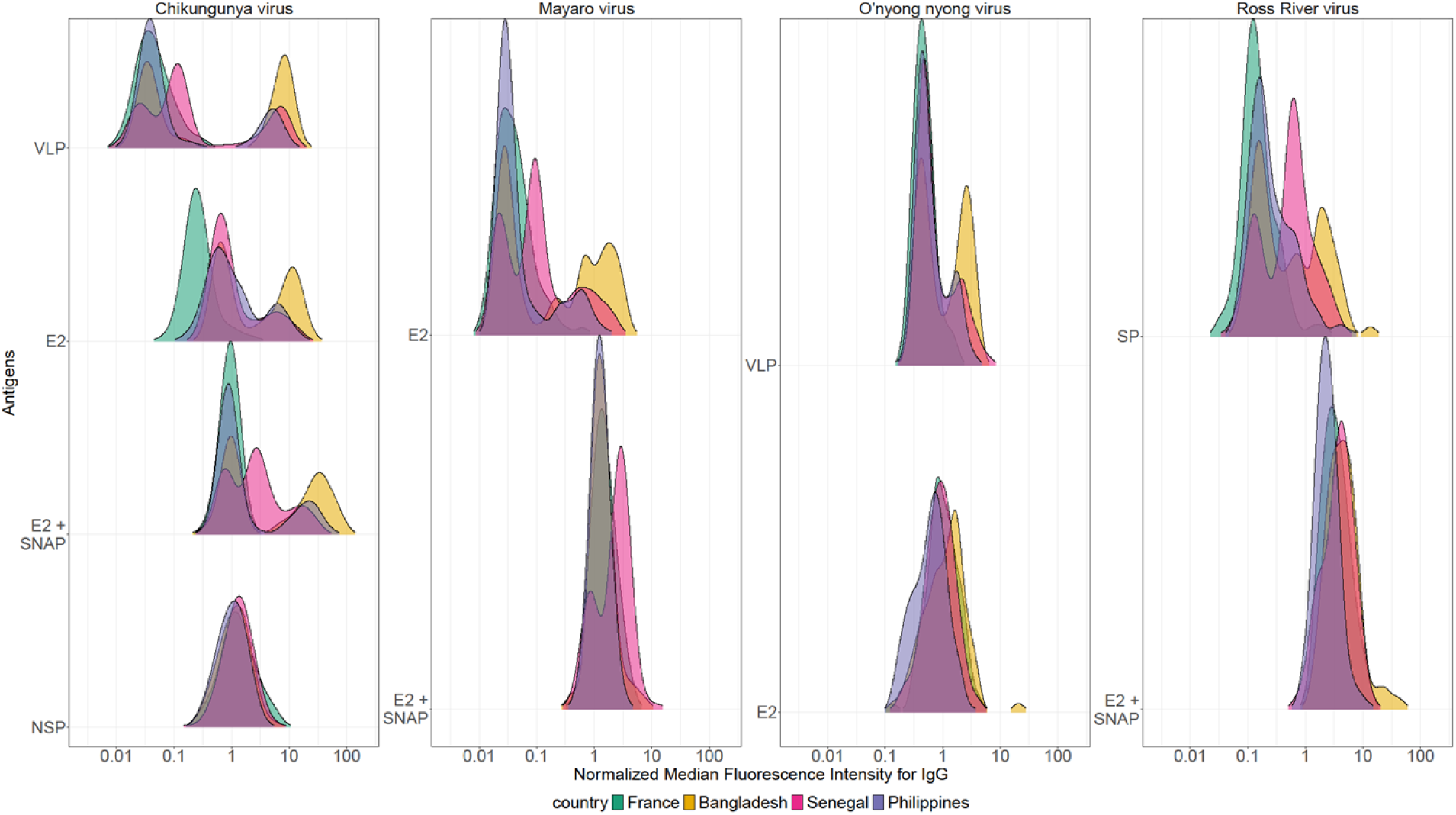
Distribution of IgG antibody responses for alphaviruses. Normalized Median Fluorescence Intensity (nMFI) for IgG responses against antigens from chikungunya, Mayaro, O’nyong-nyong, and Ross River viruses in samples from France (green), Bangladesh (yellow), Senegal (pink), and the Philippines (purple). nMFI values are derived from median fluorescence intensity (MFI) using a Luminex INTELLIFLEX assay. The antigens assessed include virus-like particles (VLP), envelope glycoprotein E2 (E2), SNAP-tagged E2 (E2 + SNAP), non-structural protein (NSP) for chikungunya virus, and structural polyprotein (SP) for Ross River virus.

A strong chikungunya IgG response was observed in samples from Bangladesh, Senegal and the Philippines, particularly against VLP, E2 and E2 + SNAP antigens. The presence of a clear bimodal distribution suggests widespread prior exposure in these regions. In contrast, samples from France displayed low IgG responses across all CHIKV antigens, consistent with limited and localized CHIKV transmission reported in southern France(28,29).

There is no clear signal for CHIKV NSP suggesting limited utility for serodiagnosis in this context. Concerning MAYV, ONNV and RRV viruses there was evidence of bi-modal distributions for MAYV E2, ONNV VLP and RR SP, but there is currently no publish evidence of these viruses circulating in Bangladesh, the Philippines or Senegal. This could be explained by cross-reactivity with other circulating alphaviruses such as CHIKV.

### Distribution of IgG antibody for flaviviruses accross four countries

To assess serological responses to orthoflaviviruses we analyzed IgG antibody levels against nine orthoflaviviruses across four countries (Figure 2). We observed a strong dengue IgG responses in samples from Bangladesh, Senegal and the Philippines, particularly against the non structural protein 1 (NS1) and virus-like particles (VLP) with evidence of bimodal distributions. In contrast, samples from France showed lower IgG levels, consistent with the near absence of endemic DENV tranmission. EDIII and EDIII + SNAP signals showed a smaller dynamic range compared to VLP and NS1.

**Figure 2:**
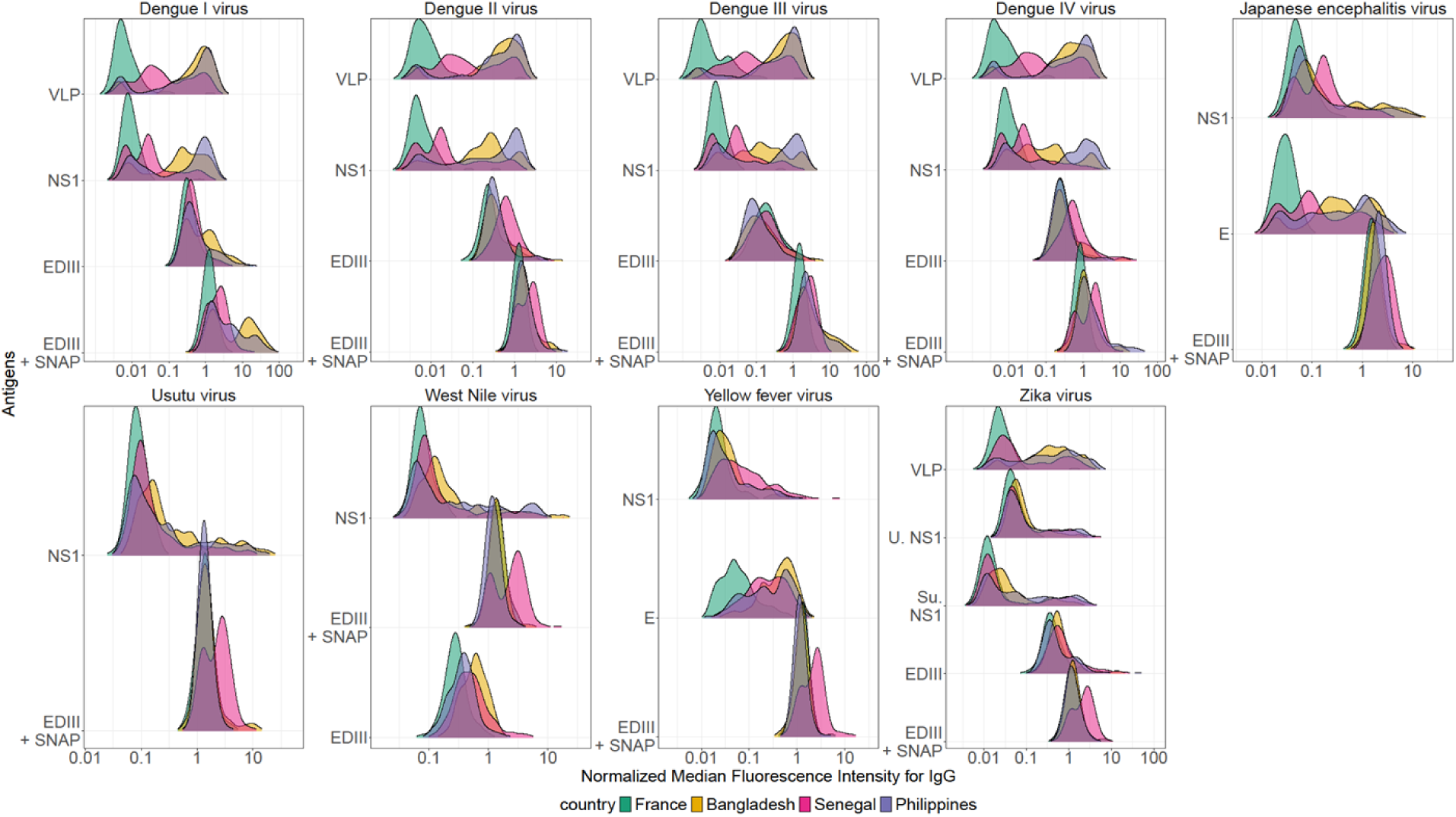
Distribution of IgG antibody responses for orthoflaviviruses. Normalized Median Fluorescence Intensity (nMFI) for IgG responses against antigens from dengue (serotypes 1-4), Japanese encephalitis, usutu, West-Nile, yellow fever and Zika viruses in samples from France (green), Bangladesh (yellow), Senegal (pink) and the Philippines (purple). nMFI values are derived from median fluorescence intensity (MFI) using a Luminex INTELLIFLEX assay. The antigens assessed include virus-like particles (VLP), non-structral protein (NS1), envelope (E), envelope domain III (EDIII), SNAP-tagged envelope domain III (EDIII + SNAP), and NS1 proteins from Uganda strains (U. NS1) and Suriname strain (Su. NS1) for Zika virus.

### Antibody kinetics following chikungunya infection

We analyzed samples from a study conducted in the Philippines where individuals were followed longitudinally for up to 3 years after RT-PCR confirmation of infection with arboviruses including CHIKV, ZIKV, and DENV1 to 4 (Supplementary Figures). Firstly, a notable increase in antibody levels is observed post-infection. CHIKV-specific antigens are highly immunogenic, leading to strong IgG responses with large boosts in antibody levels (Figure 3). Secondly, antibody responses after CHIKV infection are long-lived responses especially for VLP, E2 and E2 + SNAP. CHIKV infection caused substantial boosting of IgG responses against other alphaviruses, providing strong evidence of cross reactivity.

**Figure 3:**
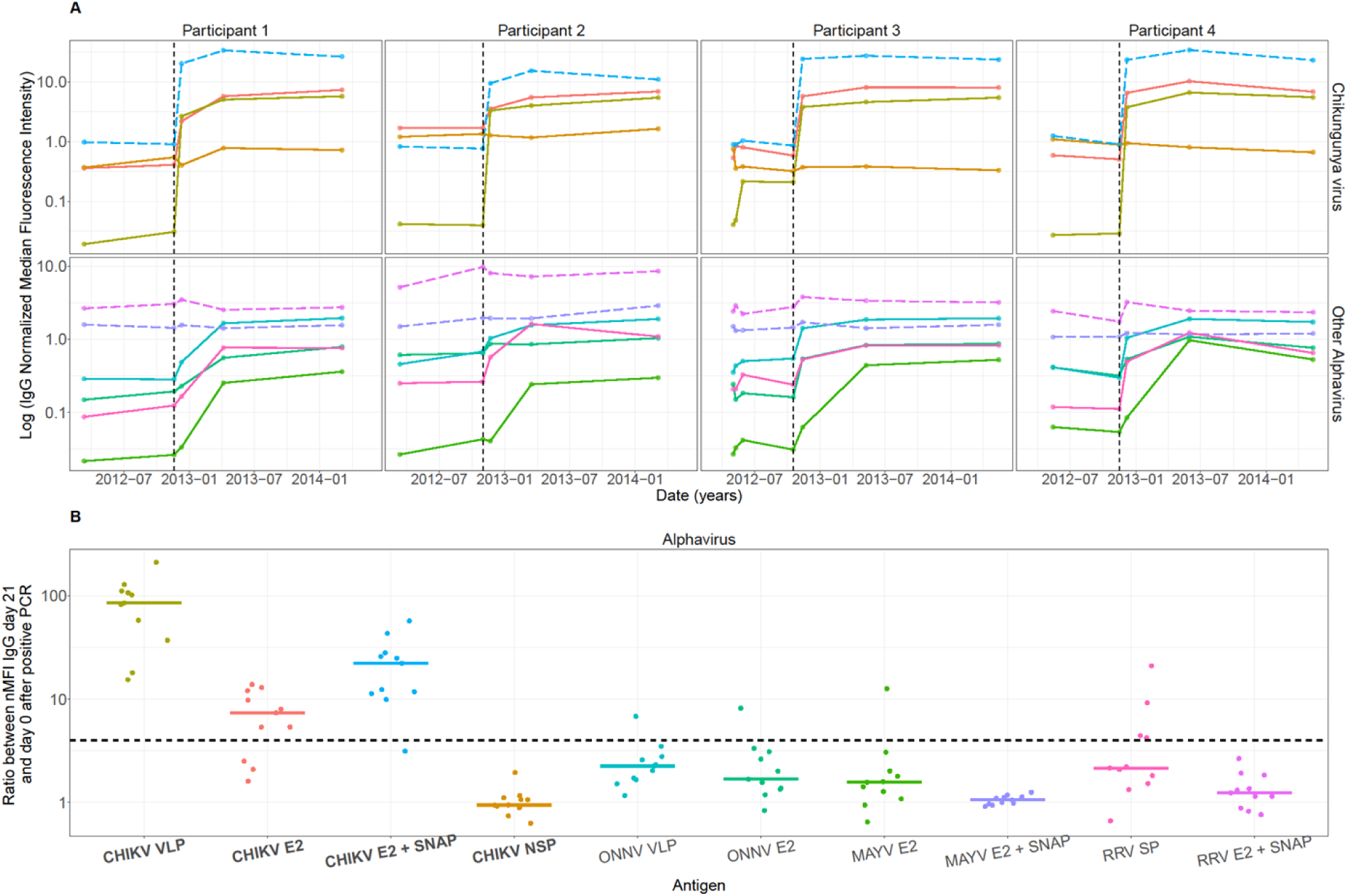
Alphavirus antibody kinetics in individuals with RT-PCR-confirmed CHIKV infection. **(A)** Longitudinal IgG antibody responses to CHIKV and other alphaviruses in 4/12 representative infected individuals. Each column represents an infected individual (participants 1 to 4), with the upper row showing responses to CHIKV-specific antigens (NSP, E2, VLP, and SNAP + E2) and the lower row displaying responses to other alphavirus antigens. The dashed vertical line indicates the date of RT-PCR-confirmed CHIKV infection. Antigens represented with dashed lines correspond to SNAP antigens. **(B)** Alphavirus antibody boost in RT-PCR-confirmed CHIKV infections. The nMFI IgG ratio between day 21 and day 0 after RT-PCR-confirmed CHIKV infection is presented. The dashed dotted line represents a 4-fold increase in nMFI. Each antigen is represented by a unique colour, defined in panel B.

### Antibody kinetics following DENV1 virus infection

Figure 4 presents the longitudinal IgG antibody responses to DENV and other orthoflavivirus antigens in four DENV1 RT-PCR-positive participants. In all individuals, a marked and rapid increase in IgG levels was observed against DENV1-specific antigens following infection, indicating a robust humoral immune response. IgG responses increased after infection and were followed by either sustained high levels or a gradual decline over time.

**Figure 4.**
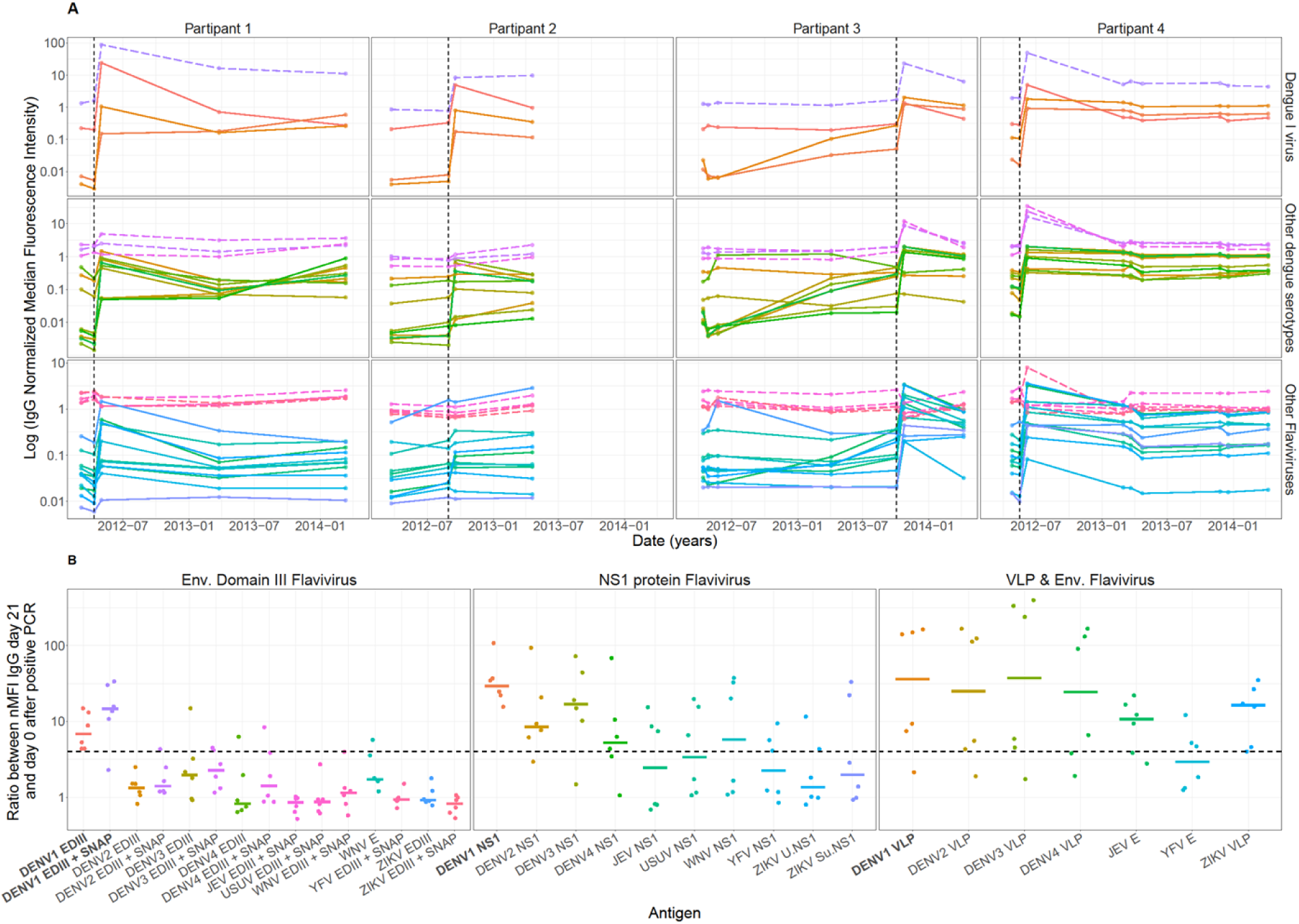
Orthoflavivirus antibody kinetics in individuals with RT-PCR-confirmed DENV1 infection. **(A)** Longitudinal IgG antibody responses to DENV1 and other orthoflavivirus in 4/12 representative infected individuals. Each column represents an infected individual (participants 1 to 4), with the top row showing responses to DENV1-specific antigens (VLP, NS1, EDIII and EDIII + SNAP), the second row showing responses to DENV2,3,4 specific antigens, and the third row displaying responses to other orthoflavivirus antigens. The dashed vertical line indicates the date of RT-PCR-confirmed DENV1 infection. Antigens represented with dashed lines correspond to SNAP antigens. **(B)** Orthoflavivirus antibody boost following RT-PCR-confirmed DENV1 infection. The nMFI IgG ratio between day 21 and day 0 after RT-PCR-confirmed DENV1 infection is presented. The dashed dotted line represents a 4-fold increase in nMFI. Each antigen is represented by a unique colour, consistent across all panels.

DENV1-infection also elicited IgG responses against other orthoflaviviruses, providing evidence of cross reactivity. This was most notable among heterologous DENV serotypes, but evident also with other orthoflaviviruses such as ZIKV. These findings underscore the complexity of orthoflavivirus serological responses. Due to this cross-reactive boosting, accurately identifying the infecting serotype based solely on IgG kinetics remains a notable challenge in endemic areas.

### Comparison of the multiplex serological assay with the Plaque Reduction Neutralisation Test (PRNT) for CHIKV

To compare the serological multiplex assay with the gold standard PRNT for CHIKV, the the association between IgG levels and PRNT titers in samples from Bangladesh was performed. First, we assessed the correlation between PRNT titers and IgG responses to individual antigens (Figure 5).

**Figure 5:**
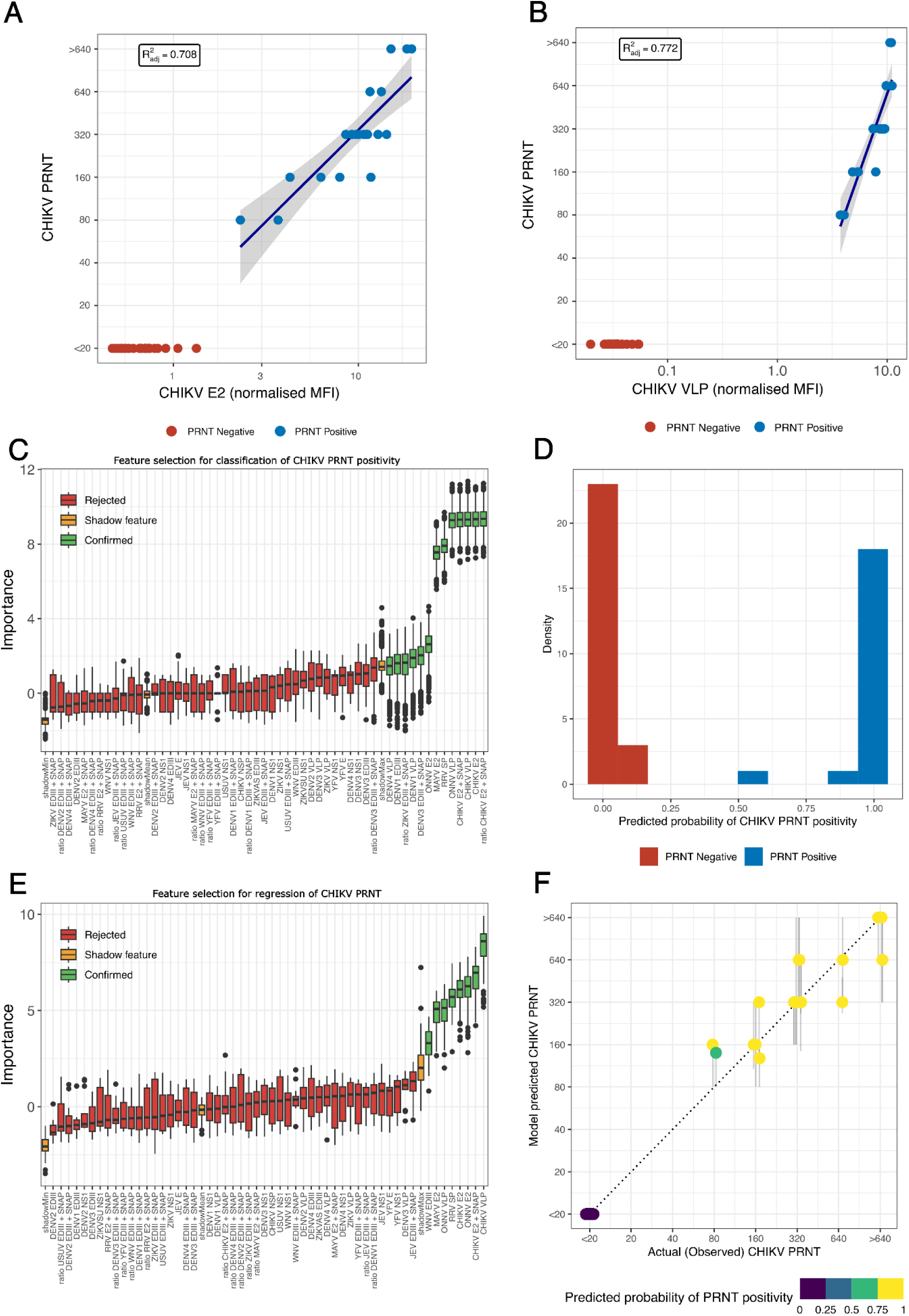
Two-step Random Forest model to predict CHIKV PRNT titer in Bangladesh samples. Comparison of CHIKV PRNT and **(A)** CHIKV E2 (nMFI) and **(B)** CHIKV VLP (nMFI). **(C)** Boruta variable importance for classification of CHIKV PRNT positivity (Step 1). The boxplots represent the distribution of importance scores for the tested antigens. Green boxes represent the features/antibody responses that contributed significantly to classification and were retained for the final classifier. Red boxes represent rejected features that did not provide meaningful classification power. Yellow boxes represent shadow features that serve as controls to determine the threshold for importance. **(D)** Distribution of the OOB predicted probability of CHIKV PRNT positivity from the random forest classification step (Step 1). PRNT positive samples are shown in blue and PRNT negative samples are shown in red. **(E)** Boruta variable importance for regression of CHIKV PRNT titers (Step 2). Green boxes represent antibody responses that were retained for the final regression model. **(F)** Aggregated OOB predictions of CHIKV PRNT titers from the full two-step random forest model (Step 1: classification and Step 2: regression). This figure compares the actual (observed) PRNT titers with model-predicted PRNT titers. Each point represents a sample. Colors indicate the predicted probability of PRNT positivity (ranging from purple [low probability] to yellow [high probability]). The dashed diagonal line represents perfect agreement between observed and predicted PRNT titer. Points clustering along this line indicate accurate predictions, while deviations suggest potential model limitations or outliers.

Figure 5A-B illustrates the strong linear relationship between IgG responses to CHIKV antigens (E2 and VLP) and PRNT titers in PRNT positive samples (adjusted R^2^ > 0.7). However, this also illustrates that a simple linear model based on the antibody response to an individual CHIKV antigen would be unable to simultaneously predict PRNT titers in positive individuals and identify PRNT-negative cases.

We therefore applied a two-step random forest model that combines classification and regression to evaluate whether a more flexible model fitted to the multiplex serological data can improve the ability to identify PRNT-negative individuals and accurately predict the PRNT titers across the full range of values (Figure 5C-F). The model showed strong performance, with most predicted titers closely matching observed titers (points clustering along the diagonal) (Figure 5F). Importantly, samples with low PRNT titers (<20) were reliably identified as negative, while higher titers were accurately estimated.

### Comparison of multiplex serological assay with PRNT assay for DENV1

For DENV1, there was a modest correlation between the PRNT titers and IgG levels to DENV NS1 and VLP antigens in PRNT positive individuals (Figure 6A and 6B). This highlights the challenge of predicting orthoflavivirus neutralization from antibody binding data alone. To explore whether multiplex IgG profiles could be used to predict PRNT outcomes, we applied the two-step Random Forest algorithm (Figure 6C-F). The model showed limited predictive performance both with regards to identifying PRNT positive individuals (Figure 6C and 6D) and for predicting PRNT titers in samples classified as PRNT positive (Figure 6E and 6F). These results indicate that predicting PRNT titers for orthoflaviviruses like DENV1 is more challenging, likely due to cross-reactivity and complex immune profiles of the Bangladeshi samples. However, the analysis did identify the most informative antigens at each model step, which included the ratio of DENV1 EDIII + SNAP, DENV1 VLP, DENV2 VLP, DENV4 VLP, and the ratio of DENV3 EDIII + SNAP highlighting the potential diagnostic relevance for these antigens (Figure 6C and 6E).

**Figure 6:**
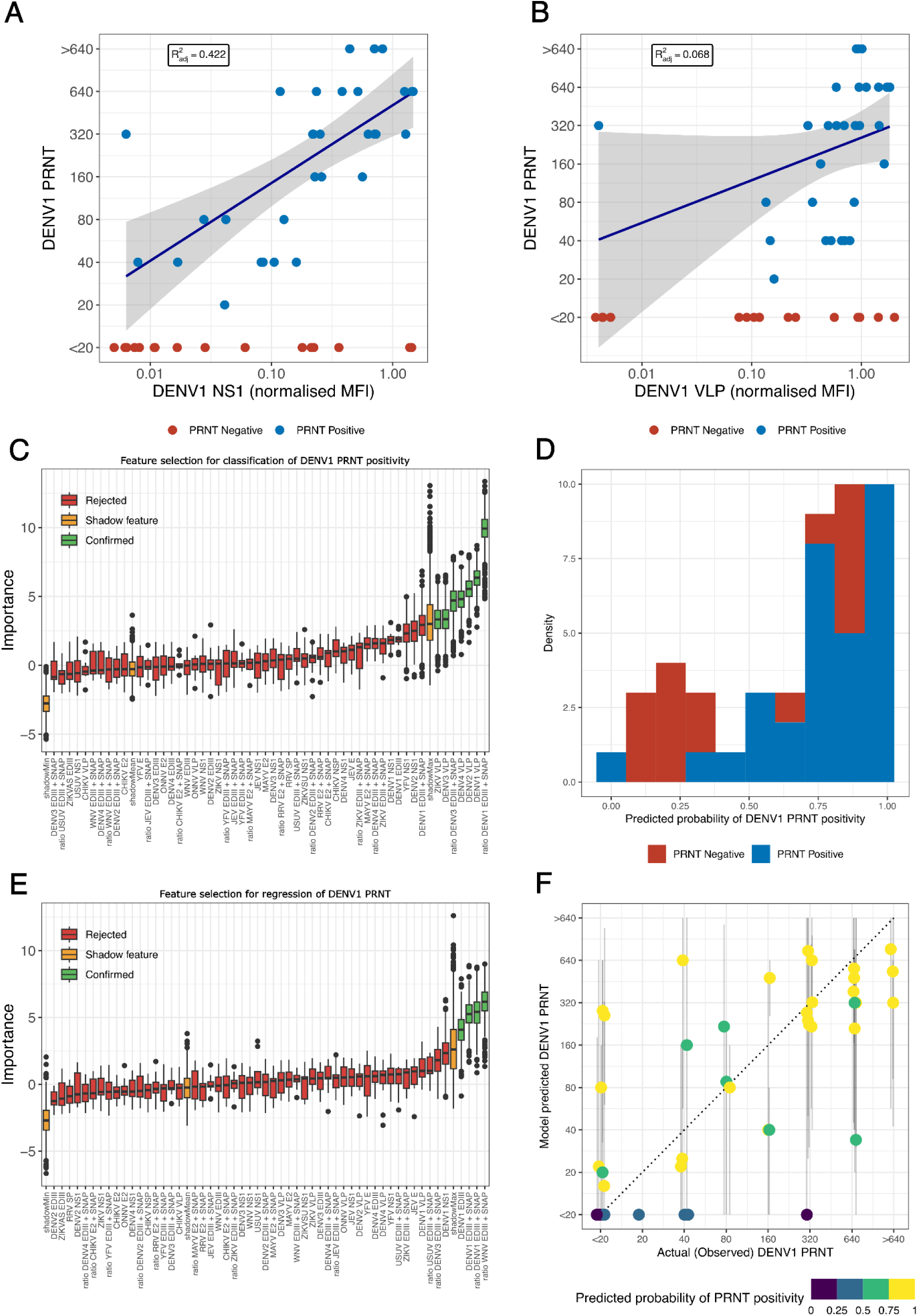
Two-step Random Forest model to predict DENV1 PRNT titer in Bangladesh samples. Comparison of DENV1 PRNT and **(A)** DENV1 NS1 (nMFI) and **(B)** DENV1 VLP (nMFI). **(C)** Boruta variable importance for classification of DENV1 PRNT positivity (Step 1). The boxplots represent the distribution of importance scores for the tested antigens. Green boxes represent the features/antibody responses that contributed significantly to classification and were retained for the final classifier. Red boxes represent rejected features that did not provide meaningful classification power. Yellow boxes represent shadow features that serve as controls to determine the threshold for importance. **(D)** Distribution of the OOB predicted probability of DENV1 PRNT positivity from the random forest classification step (Step 1). PRNT positive samples are shown in blue and PRNT negative samples are shown in red. **(E)** Boruta variable importance for regression of DENV1 PRNT titers (Step 2). Green boxes represent antibody responses that were retained for the final regression model. **(F)** Aggregated OOB predictions of DENV1 PRNT titers from the full two-step random forest model (Step 1: classification and Step 2: regression). This figure compares the actual (observed) PRNT titers with model-predicted PRNT titers. Each point represents a sample. Colors indicate the predicted probability of PRNT positivity (ranging from purple [low probability] to yellow [high probability]). The dashed diagonal line represents perfect agreement between observed and predicted PRNT titer. Points clustering along this line indicate accurate predictions, while deviations suggest potential model limitations or outliers.

## Discussion

Serological surveillance is a key tool for the major challenge of understanding the spread and immune landscape of arboviruses, particularly in regions where multiple viruses co-circulate. In this study, we applied a high-throughput Luminex-based multiplex immunoassay to assess IgG responses to alphaviruses and orthoflaviviruses across four epidemiologically distinct countries: Bangladesh, the Philippines, Senegal, and France.

Our findings highlight both the potential and limitations of IgG-based serology assays in distinguishing infections with closely related arboviruses, especially given the known cross-reactivity within viral families. IgG is a valuable marker for assessing past exposure, as it typically becomes detectable within a few weeks after infection.(30) Comparaison of traditional methods with our multiplex bead-based immunoassay revealed substantial advantages such as simultaneous detection of IgG antibodies against 44 different antigens using a single, small-volume serum sample at low cost conserving high specificity and efficiency. By measuring responses to a panel of arbovirus antigens across multiple geographic settings, and testing both longitudinal samples from RT-PCR–confirmed infections and cross-sectional samples (including those with paired neutralization data), we identified markers with good discriminatory power between seropositive and seronegative individuals.

A strong chikungunya IgG response was observed in samples from Bangladesh and Senegal, particularly against VLP, E2, and E2 + SNAP antigens. The presence of a clear bimodal distribution suggests widespread prior exposure in these regions. In contrast, samples from France displayed low IgG responses across all CHIKV antigens, consistent with limited and localized CHIKV transmission reported in southern France(28,29). The distribution of the IgG response to CHIKV antigens exhibited a clear peak corresponding to infected individuals. For the uninfected individuals, there was substantial variation between populations which we attribute to differences in total IgG levels between populations(31),(32). Similar patterns were observed in the IgG kinetics of RT-PCR–confirmed CHIKV infections, with VLP, E2, and E2 + SNAP responses showing a marked post-infection increase compared with other alphavirus antigens (Figure 3). These antigens show strong potential as markers for distinguishing alphavirus infections. In contrast, responses to the CHIKV NSP antigen were less discriminative, indicating limited diagnostic utility (Figure 1 and 3).

In Senegal, where low-level The transmission of ONNV is restricted to Africa(33). However, the distribution of IgG responses to ONNV was similar between Senegal, where trasmission has been documented^33^, and Bangladesh and the Philippines, where circulation is not documented. The ONNV VLP antigen showed a response profile similar to CHIKV VLP, suggesting cross-reactivity and limited specificity. In contrast, the ONNV E2 antigen elicited low and consistent IgG levels across all four countries, indicating minimal exposure, and supporting its potential as a specific marker for ONNV seroprevalence. Despite the close phylogenetic relationship between ONNV and CHIKV, cross-reactivity after CHIKV infection was modest and comparable to that observed with MAYV, a more distantly related alphavirus(34) (Figure 3). This was unexpected, and although a slight boost in IgG was observed for ONNV VLP, ONNV E2, MAYV E2 and MAYV E2 + SNAP following CHIKV infection, these antigens still performed well to distinguishing CHIKV from other alphavirus infections. The inclusion of longitudinal samples from RT-PCR–confirmed cases was critical for identifying the most informative markers, as it enabled evaluation of antigen responses over time, something not possible in cross-sectional studies where past exposure is often uncertain.

Although RRV has not been officially reported in Senegal, France, or Bangladesh, the RRV SP antigen elicited strong IgG responses in participants from Bangladesh. This may reflect cross-reactivity with other circulating alphaviruses, particularly CHIKV, as evidenced by the boost in anti-RRV SP antibodies following RT-PCR-confirmed CHIKV infection (Figure 3). Notably, recent studies suggest possible RRV circulation in parts of the Pacific, highlighting the need for further investigation into its geographic distribution(35,36). The SNAP-tagged E2 antigen for RRV produced uniformly low IgG signals, highlighting its potential for improved specificity in serodiagnosis. In contrast, for MAYV which is endemic primarily to South and Central America(37), IgG responses to the E2 protein in non-endemic regions likely reflect cross-reactivity rather than true exposure (Figure 1). Again, the SNAP-tagged E2 antigen showed lower IgG responses. This pattern was also evident in Figure 3, where CHIKV infection elicited a boost in IgG against MAYV E2 and MAYV E2 + SNAP, but these increases were less pronounced than those observed for CHIKV antigens, reinforcing its value for reducing cross-reactivity in serological assays(38).

Orthoflavivirus IgG responses are known to exhibit substantial cross-reactivity, complicating serological differentiation. Among DENV antigens, NS1 and VLP consistently generated the highest IgG levels, indicating their sensitivity for identifying past exposure. However due to the high degree of correlation of these antibody responses between different orthoflaviviruses, these antigens have limited specificty. Following RT-PCR-confirmed DENV1 infection, the boost to anti-EDIII antibodies was notably smaller than for the VLP and NS1 antigens, indicating that EDIII is a less immunogenic biomarker of infection. However, in contrast to the VLP and NS1 antigens, the boost in anti-EDIII antibodies following infection was much more pronouced for the infecting DENV serotype than for non-infecting serotypes or related orthoflaviviruses, supporting the hypothesis that anti-EDIII antibodies provide greater specificity (Figure 4). SNAP-tagged EDIII antigens displayed similar patterns to untagged EDIII but with potential for enhanced sensitivity and specificity. Unlike non-SNAP-tagged antigens, which bind covalently to beads without controlled orientation, SNAP-tagged antigens are specifically coupled via the SNAP domain using BG-PEG-NH_2_ substrate, leaving the viral antigen fully exposed for antibody recognition and reducing background cross-reactivity.

Responses to JEV provided further evidence of orthoflavivirus cross-reactivity. In Senegal, where no JEV circulation has been reported, strong IgG responses against E and NS1 were nonetheless detected, suggesting cross-reactivity with other orthoflaviviruses. In Bangladesh, where JEV is known to circulate(17), strong IgG responses against E and NS1 were also observed. However, the lack of corresponding responses to SNAP-tagged EDIII indicates that much of this reactivity may reflect cross-reactivity with other orthoflaviviruses rather than JEV-specific infection. For both USUV and WNV, interpretation of IgG responses in non-endemic regions was complicated by cross-reactivity. Although no USUV circulation has been documented in Bangladesh or the Philippines, high NS1-specific IgG levels were observed. Similarly, WNV-specific responses were elevated in all sites despite confirmed circulation only in Senegal(39). In both cases, SNAP-tagged EDIII antigens elicited low IgG signals in non-endemic regions, with a bimodal distribution in Senegal suggestive of true WNV exposure. These findings support the use of engineered envelope antigens for improved specificity.

ZIKV circulation has been documented in both Africa and Asia, but with notable regional differences. In Senegal where ZIKV circulation has been well documented^42^ and in Bangladesh, where circulation appears limited, with a single confirmed case reported in 2016(42) and a cluster of five cases identified in 2023(43), strong IgG responses against VLP, U.NS1, and Su.NS1 antigens were observed. In contrast, responses to EDIII and SNAP-tagged EDIII antigens were detected exclusively in Senegalese samples, consistent with true ZIKV exposure in this setting and supporting the interpretation that reactivity observed in Bangladesh may not reflect true ZIKV circulation. Similarly, YFV circulation has been well documented in West Africa, including Senegal, where serological surveys, vector studies, and recurrent outbreaks confirm endemic transmission(44) and also vaccination campaigns have occurred, whereas in Bangladesh no evidence of YFV circulation has been reported. Indeed, we observed high IgG responses against NS1 and E antigens in samples from both regions. However, responses to SNAP-tagged EDIII antigen were detected only in Senegalese samples, consistent with true YFV exposure or vaccination responses. At present, our in-house multiplex bead-based immunoassay does not allow us to discriminate between antibody responses induced by natural YFV infection and those generated by vaccination.

IgG responses following DENV1 infection (Figure 4A) were less pronounced than those observed for CHIKV, likely due to broader cross-reactivity among orthoflaviviruses. In the Philippines, co-circulation(45) of DENV, ZIKV(46), and JEV(47) complicates clinical diagnosis and public health responses.

In Figure 4B, increases in IgG levels against multiple orthoflavivirus antigens indicate that DENV1 EDIII and SNAP-tagged EDIII provide improved discrimination between orthoflavivirus infections. In contrast, NS1 and VLPs exhibited higher cross-reactivity. Although IgG-based assays are generally affected by cross-reactivity due to affinity maturation and class switching, a similar pattern was observed for both IgM and IgA responses (Supplementary Figures S1-6). This likely reflects the high degree of structural similarity among orthoflavivirus antigens, particularly in conserved regions of NS1 and the envelope protein. Early polyclonal B cell activation, especially during secondary or sequential flavivirus infections, may also contribute to cross-reactive IgM and IgA production(48). The consistently elevated cross-reactivity observed with VLPs antigens across all isotypes highlights the role of epitope conservation in shaping the humoral immune response.

To investigate the association between serological profiles and functional immunity, a two-step Random Forest approach to predict PRNT-based neutralizing antibody responses using serological data was applied. Our findings underscore that the strength of serology as a surrogate for neutralization depends heavily on the virus and background levels of cross-reactive immunity. For CHIKV, a strong correlation between IgG responses, particularly to E2 and VLP antigens, and PRNT titers were observed, confirming that our in-house serological assay reliably reflects functional antibody activity. Both classification and regression models performed robustly, accurately identifying seropositive individuals and quantitatively predicting PRNT titers. These results support that the multiplex immunoassay data may serve as a reliable surrogate for PRNT in CHIKV serodiagnostics, enabling both classification and quantitative prediction of neutralizing capacity.

In contrast, predictive performance was substantially lower for DENV1, where IgG-based models struggled to accurately estimate neutralization. This disparity likely reflects the high degree of cross-reactivity and complex patterns of immunodominance and original antigenic sin among co-circulating orthoflaviviruses, which disrupts the relationship between binding and neutralizing antibodies. These results highlight that while machine learning tools can enhance the utility of multiplex serology, their predictive power depends on a clear immunological signal. Going forward, integrating serology with machine learning holds promise for scalable immunosurveillance, provided antigen selection and immunological context are carefully considered.

Our findings highlight the widespread issue of cross-reactivity among alphaviruses and orthoflaviviruses in serological assays and the resulting challenges for accurate serosurveillance. The use of SNAP-tagged EDIII and E2 antigens demonstrated improved specificity across multiple virus targets, emphasizing the importance of antigen engineering and the strategic use of multiple antigens in the development of reliable serosurveillance tools. Regional differences in IgG profiles further underscore the need for context-specific validation of serological markers, especially in areas with overlapping arbovirus transmission. Our longitudinal approach, in contrast to cross-sectional data (Figure 1), reveals the temporal dynamics of IgG responses and strengthens the ability to distinguish true infection from background cross-reactivity. Our study has several limitations that should be acknowledged. Most notably, we did not have access to commercially available reference samples or larger number of samples with PCR-confirmed arbovirus infections, which limited our ability to fully validate the assay and include standardized positive and negative controls. Despite these limits, our results provide valuable insights into arbovirus serology across regions and highlight the importance of future studies incorporating larger, well-defined cohorts and reference panels.

## Author Contribution

LG, FD and MFG performed the multiplex serological assays.

LC, DH and PhD provided the in-house recombinant proteins.

IKY, TH, EG, NV, SW and MZR provided the PRNT data.

IVW, MN, JCM, IL, CD and HS were involved in study design, patient recruitment, supervision, sample collection and distribution.

LG, VY, MW and JV analysed the data.

LG wrote the manuscript with input from MW and JV. All authors contributed to the review and editing of the manuscript. All authors have read and agreed to the published version of the manuscript.

## Disclaimer

Material has been reviewed by the Walter Reed Army Institute of Research. There is no objection to its presentation and/or publication. The opinions or assertions contained herein are the private views of the author, and are not to be construed as official, or as reflecting true views of the Department of the Army or the Department of Defense. The investigators have adhered to the policies for protection of human subjects as prescribed in AR 70–25.

## Data Availability

The data that support the findings of this study are openly available in the Supplementary information.

## Acknolegments

Maylis Douine and Alice Sanna are thanked for sharing a pool of serum from arbovirus-exposed individuals.

## References

1. Madewell ZJ. Arboviruses and Their Vectors. South Med J. oct 2020;113(10):520–3.

2. Dengue worldwide overview [Internet]. 2024 [cité 17 janv 2025]. Disponible sur: https://www.ecdc.europa.eu/en/dengue-monthly

3. Chikungunya worldwide overview [Internet]. 2024 [cité 17 janv 2025]. Disponible sur: https://www.ecdc.europa.eu/en/chikungunya-monthly

4. Samarasekera U, Triunfol M. Concern over Zika virus grips the world. The Lancet. 6 févr 2016;387(10018):521–4.

5. Campos GS, Bandeira AC, Sardi SI. Zika Virus Outbreak, Bahia, Brazil. Emerg Infect Dis. oct 2015;21(10):1885–6.

6. Zardini A, Menegale F, Gobbi A, Manica M, Guzzetta G, d’Andrea V, et al. Estimating the potential risk of transmission of arboviruses in the Americas and Europe: a modelling study. Lancet Planet Health. 1 janv 2024;8(1):e30–40.

7. Fritzell C, Rousset D, Adde A, Kazanji M, Kerkhove MDV, Flamand C. Current challenges and implications for dengue, chikungunya and Zika seroprevalence studies worldwide: A scoping review. PLoS Negl Trop Dis. 16 juill 2018;12(7):e0006533.

8. Vatti A, Monsalve DM, Pacheco Y, Chang C, Anaya JM, Gershwin ME. Original antigenic sin: A comprehensive review. J Autoimmun. sept 2017;83:12–21.

9. Ding X, Zhao F, Liu Z, Yao J, Yu H, Zhang X. Original antigenic sin: A potential double-edged effect for vaccine improvement. Biomed Pharmacother. 1 sept 2024;178:117187.

10. Endale A, Medhin G, Darfiro K, Kebede N, Legesse M. Magnitude of Antibody Cross-Reactivity in Medically Important Mosquito-Borne Flaviviruses: A Systematic Review. Infect Drug Resist. 19 oct 2021;14:4291–9.

11. Varghese J, De Silva I, Millar DS. Latest Advances in Arbovirus Diagnostics. Microorganisms. 28 avr 2023;11(5):1159.

12. Gil-Mora J, Acevedo-Gutiérrez LY, Betancourt-Ruiz PL, Martínez-Diaz HC, Fernández D, Bopp NE, et al. Arbovirus Antibody Seroprevalence in the Human Population from Cauca, Colombia. Am J Trop Med Hyg. déc 2022;107(6):1218–25.

13. Fischer C, Jo WK, Haage V, Moreira-Soto A, de Oliveira Filho EF, Drexler JF. Challenges towards serologic diagnostics of emerging arboviruses. Clin Microbiol Infect Off Publ Eur Soc Clin Microbiol Infect Dis. sept 2021;27(9):1221–9.

14. Hein LD, Castillo IN, Medina FA, Vila F, Segovia-Chumbez B, Muñoz-Jordán JL, et al. Multiplex sample-sparing assay for detecting type-specific antibodies to Zika and dengue viruses: an assay development and validation study. Lancet Microbe. 1 févr 2025;6(2):100951.

15. Salje H, Cauchemez S, Alera MT, Rodriguez-Barraquer I, Thaisomboonsuk B, Srikiatkhachorn A, et al. Reconstruction of 60 Years of Chikungunya Epidemiology in the Philippines Demonstrates Episodic and Focal Transmission. J Infect Dis. 15 févr 2016;213(4):604–10.

16. Alera MT, Srikiatkhachorn A, Velasco JM, Tac-An IA, Lago CB, Clapham HE, et al. Incidence of Dengue Virus Infection in Adults and Children in a Prospective Longitudinal Cohort in the Philippines. PLoS Negl Trop Dis. 4 févr 2016;10(2):e0004337.

17. Duque MP, Naser AM, dos Santos GR, O’Driscoll M, Paul KK, Rahman M, et al. Informing an investment case for Japanese encephalitis vaccine introduction in Bangladesh. Sci Adv. 9 août 2024;10(32):eadp1657.

18. Niang M, Sandfort M, Mbodj AF, Diouf B, Talla C, Faye J, et al. Fine-scale Spatiotemporal Mapping of Asymptomatic and Clinical Plasmodium falciparum Infections: Epidemiological Evidence for Targeted Malaria Elimination Interventions. Clin Infect Dis Off Publ Infect Dis Soc Am. 16 déc 2021;73(12):2175–83.

19. Woudenberg T, Pinaud L, Garcia L, Tondeur L, Pelleau S, De Thoisy A, et al. Estimated protection against COVID-19 based on predicted neutralisation titres from multiple antibody measurements in a longitudinal cohort, France, April 2020 to November 2021. Eurosurveillance. 22 juin 2023;28(25):2200681.

20. Beck C, Desprès P, Paulous S, Vanhomwegen J, Lowenski S, Nowotny N, et al. A High-Performance Multiplex Immunoassay for Serodiagnosis of Flavivirus-Associated Neurological Diseases in Horses. BioMed Res Int. 2015;2015:678084.

21. Bloch E, Baudemont G, Donnadieu F, Garcia L, Pelleau S, Consortium SS, et al. Investigation of the sero-epidemiology of vaccine preventable diseases and common viral infections in French populations using a multiplex serological assay [Internet]. medRxiv; 2024 [cité 20 août 2025]. p. 2024.04.26.24306413. Disponible sur: https://www.medrxiv.org/content/10.1101/2024.04.26.24306413v1

22. Bloch E, Baudemont G, Donnadieu F, Garcia L, Pelleau S, Consortium SS, et al. Investigation of the sero-epidemiology of vaccine preventable diseases and common viral infections in French populations using a multiplex serological assay [Internet]. medRxiv; 2024 [cité 4 juill 2025]. p. 2024.04.26.24306413. Disponible sur: https://www.medrxiv.org/content/10.1101/2024.04.26.24306413v1

23. Russell PK, Nisalak A, Sukhavachana P, Vivona S. A plaque reduction test for dengue virus neutralizing antibodies. J Immunol Baltim Md 1950. août 1967;99(2):285–90.

24. Chusri S, Siripaitoon P, Silpapojakul K, Hortiwakul T, Charernmak B, Chinnawirotpisan P, et al. Kinetics of Chikungunya Infections during an Outbreak in Southern Thailand, 2008–2009. 5 mars 2014 [cité 3 mars 2025]; Disponible sur: https://www.ajtmh.org/view/journals/tpmd/90/3/article-p410.xml

25. Levitt NH, Ramsburg HH, Hasty SE, Repik PM, Cole FE, Lupton HW. Development of an attenuated strain of chikungunya virus for use in vaccine production. Vaccine. 1 sept 1986;4(3):157–62.

26. Vasilakis N, Durbin AP, da Rosa APAT, Munoz-Jordan JL, Tesh RB, Weaver SC. Antigenic relationships between sylvatic and endemic dengue viruses. Am J Trop Med Hyg. juill 2008;79(1):128–32.

27. Kursa MB, Rudnicki WR. Feature Selection with the Boruta Package. J Stat Softw. 16 sept 2010;36:1–13.

28. Grandadam M, Caro V, Plumet S, Thiberge JM, Souarès Y, Failloux AB, et al. Chikungunya virus, southeastern France. Emerg Infect Dis. mai 2011;17(5):910–3.

29. Calba C, Guerbois-Galla M, Franke F, Jeannin C, Auzet-Caillaud M, Grard G, et al. Preliminary report of an autochthonous chikungunya outbreak in France, July to September 2017. Euro Surveill Bull Eur Sur Mal Transm Eur Commun Dis Bull. sept 2017;22(39):17–00647.

30. Wu Q, Jing Q, Wang X, Yang L, Li Y, Chen Z, et al. Kinetics of IgG Antibodies in Previous Cases of Dengue Fever—A Longitudinal Serological Survey. Int J Environ Res Public Health. janv 2020;17(18):6580.

31. Harkness T, Fu X, Zhang Y, Choi HK, Stone JH, Blumenthal KG, et al. Immunoglobulin G and immunoglobulin G subclass concentrations differ according to sex and race. Ann Allergy Asthma Immunol Off Publ Am Coll Allergy Asthma Immunol. août 2020;125(2):190–195.e2.

32. Logie DE, McGREGOR IA, Rowe DS, Billewicz WZ. Plasma immunoglobulin concentrations in mothers and newborn children with special reference to placental malaria.

33. Mutsaers M, Engdahl CS, Wilkman L, Ahlm C, Evander M, Lwande OW. Vector competence of Anopheles stephensi for O’nyong-nyong virus: a risk for global virus spread. Parasit Vectors. 17 avr 2023;16(1):133.

34. Bessaud M, Peyrefitte CN, Pastorino BAM, Gravier P, Tock F, Boete F, et al. O’nyong-nyong Virus, Chad - Volume 12, Number 8—August 2006 - Emerging Infectious Diseases journal - CDC. [cité 14 août 2025]; Disponible sur: https://wwwnc.cdc.gov/eid/article/12/8/06-0199_article

35. Lau C, Aubry M, Musso D, Teissier A, Paulous S, Desprès P, et al. New evidence for endemic circulation of Ross River virus in the Pacific Islands and the potential for emergence. Int J Infect Dis IJID Off Publ Int Soc Infect Dis. avr 2017;57:73–6.

36. Aubry M, Teissier A, Huart M, Merceron S, Vanhomwegen J, Roche C, et al. Ross River Virus Seroprevalence, French Polynesia, 2014-2015. Emerg Infect Dis. oct 2017;23(10):1751–3.

37. Mayaro virus in Latin America and the Caribbean [Internet]. Pan American Journal of Public Health; [cité 14 mai 2025]. Disponible sur: https://journal.paho.org/en/articles/mayaro-virus-latin-america-and-caribbean

38. Hozé N, Salje H, Rousset D, Fritzell C, Vanhomwegen J, Bailly S, et al. Reconstructing Mayaro virus circulation in French Guiana shows frequent spillovers. Nat Commun. 5 juin 2020;11(1):2842.

39. Ndione MHD, Ndiaye EH, Faye M, Diagne MM, Diallo D, Diallo A, et al. Re-Introduction of West Nile Virus Lineage 1 in Senegal from Europe and Subsequent Circulation in Human and Mosquito Populations between 2012 and 2021. Viruses. 6 déc 2022;14(12):2720.

40. Diallo D, Sall AA, Diagne CT, Faye O, Faye O, Ba Y, et al. Zika Virus Emergence in Mosquitoes in Southeastern Senegal, 2011. PLoS ONE. 13 oct 2014;9(10):e109442.

41. Hossain MdG, Nazir KHMNH, Saha S, Rahman MdT. Zika virus: A possible emerging threat for Bangladesh! J Adv Vet Anim Res. 10 nov 2019;6(4):575–82.

42. Muraduzzaman AKM, Sultana S, Shirin T, Khatun S, Islam M, Rahman M. Introduction of Zika virus in Bangladesh: An impending public health threat. Asian Pac J Trop Med. sept 2017;10(9):925–8.

43. Hasan A, Hossain MM, Zamil MF, Trina AT, Hossain MS, Kumkum A, et al. Concurrent transmission of Zika virus during the 2023 dengue outbreak in Dhaka, Bangladesh. PLoS Negl Trop Dis. 30 janv 2025;19(1):e0012866.

44. Gallon S, Sy M, Tonto PB, Ndiaye IM, Toure M, Gaye A, et al. Seropositivity to Dengue, Zika, Yellow Fever, and West Nile Viruses in Senegal, West Africa. J Med Virol. avr 2025;97(4):e70338.

45. Daroy MLG. Zeroing on mosquito-borne viruses: Dengue virus, chikungunya virus and Japanese encephalitis virus. J Microb Biochem Technol [Internet]. [cité 14 mai 2025]; Disponible sur: https://www.walshmedicalmedia.com/

46. Biggs JR, Sy AK, Brady OJ, Kucharski AJ, Funk S, Tu YH, et al. Serological Evidence of Widespread Zika Transmission across the Philippines. Viruses. août 2021;13(8):1441.

47. Nealon J, Taurel AF, Yoksan S, Moureau A, Bonaparte M, Quang LC, et al. Serological Evidence of Japanese Encephalitis Virus Circulation in Asian Children From Dengue-Endemic Countries. J Infect Dis. 1 févr 2019;219(3):375–81.

48. Nawa M, Yamada KI, Takasaki T, Akatsuka T, Kurane I. Serotype-Cross-Reactive Immunoglobulin M Responses in Dengue Virus Infections Determined by Enzyme-Linked Immunosorbent Assay. Clin Diagn Lab Immunol. sept 2000;7(5):774–7.

